# Efficacy and Tolerability of Olaparib Plus Paclitaxel in Patients with hereditary gastric cancer linked to a family history of hereditary breast and ovarian cancer (HBOC)

**DOI:** 10.1101/2024.09.03.24313047

**Authors:** Takuma Hayashi, Kenji Sano, Mako Okada, Manabu Muto, Ikuo Konishi

**Affiliations:** Cancer Medicine, National Hospital Organization Kyoto Medical Center, Kyoto 612-8555, Kyoto; Pathological Division, Shinshu University Hospital, Matsumoto, 390-0877, Nagano; Medical Oncology, Kyoto University Hospital, Kyoto 612-8555, Kyoto; Kyoto University School of Medicine, Kyoto 606-8507, Kyoto

**Author notes:** **Corresponding author:** Takuma Hayashi, Cancer Medicine, National Hospital Organization Kyoto Medical Center, Fushimi-ku, Kyoto 612-8555, Kyoto.

**Keywords:** Gastric cancer, Helicobacter pylori, BRCA1, BRCA2, HBOC

## Abstract

Helicobacter pylori (H. pylori), a type of nematode, is a common cause of chronic stomach infection around the world. In 2014, the World Health Organization (WHO) reported that H. pylori infection is a leading cause of gastric cancer (80%) worldwide and has specific carcinogenic factors. H. pylori infection is presumed to be the cause of gastric cancer in more than 98% of gastric cancer patients in East Asia, including Japan. However, only some types of gastric cancers are associated with H. pylori infection. Previous clinical studies have revealed that the bacterium secretes the cytotoxin-associated gene A (CagA) antigen, which inhibits the nuclear translocation of breast cancer susceptibility gene 1 (BRCA1) or BRCA2, a factor that repairs damaged DNA. Accordingly, an association has been pointed out between hereditary breast and ovarian cancers (HBOC) and the development of gastric cancer; however, there is a lack of clarity about the detailed mechanisms underlying the development of gastric cancer by H. pylori infection. Using the information base on hereditary cancers built up through cancer genomic medicine, our group highlighted the higher incidence of gastric cancer in HBOC families, with a preponderance for gastric cancer in male patients from HBOC families. We also verified the safety and efficacy of using poly ADP-ribose polymerase (PARP) inhibitors in patients with hereditary gastric cancer. The present study offers substantial evidence for guiding the establishment of early treatment for patients with advanced/metastatic gastric cancer in whom BRCA1/2 mutations have been detected.

## Introduction

Gastric cancer involves uncontrolled neoplastic transformation of the mucosal epithelial cells lining the stomach wall to form tumors that may recur or metastasize (malignant tumors) (1). Cancer cells proliferate and gradually invade deeper into the gastric submucosa, muscularis propria, and serosal layers. Eventually, these cancer cells may cross the serosa, seep out to spread to the surrounding areas, and invade the nearby organs, such as the large intestine, pancreas, diaphragm, and liver. In the case of advanced gastric cancer, these cancer cells can spread beyond the outer serosa and scatter throughout the abdomen, known as peritoneal dissemination. In addition, distant metastasis can occur via lymphatic or blood flow.

The typical symptoms of gastric cancer include stomach pain, discomfort, heartburn, nausea, and loss of appetite (2). Additionally, bleeding can also occurs from gastric cancer tissue, causing anemia and black stools; however, these symptoms are also reported by patients with gastritis and/or gastric ulcers. In contrast, patients with early-stage gastric cancer may not experience any symptoms at all (3), with many patients remaining asymptomatic even after the disease has progressed significantly. As there are only a few typical subjective symptoms characteristic of gastric cancer in the early stages, it is important to understand the molecular pathogenesis of gastric cancer that can be used to establish testing and treatment methods for determining the onset of gastric cancer in the initial phases.

It is presumed that cancer development occurs because of the accumulation of multiple cancer-related gene mutations within a single cell. Specific genetic pathogenic variants and driver genes have been identified for each type of cancer. Mutations in the gene for the epidermal growth factor receptor (EGFR) protein were first identified as the driver for lung cancer, which brought about a major paradigm shift in the treatment of lung cancer (4). EGFR gene mutations were detected in approximately half of the Japanese patients with lung adenocarcinomas (5). Currently, the mutated KRAS proto-oncogene (KRAS) is accepted as the most common oncogene in human cancers, accounting for approximately 30% of all solid tumors and pancreatic adenocarcinomas (the most common type of pancreatic cancer). KRAS is mutated in 95% of cases (6). Likewise, previous clinical studies have shown that H. pylori infection is associated with the development of gastric cancer and some malignant lymphomas (7).

H. pylori infection is a known cause of inflammation and ulcers in the stomach and small intestine, with 90% of all gastric cancer patients testing positive for H. pylori infection, making it a significant risk factor for the development of gastric cancer (8, 9). A previous study demonstrated that H. pylori infects gastric mucosal epithelial cells, and the cytotoxin-associated gene A (CagA) antigen inhibits the nuclear migration of breast cancer susceptibility 1 (BRCA1) and BRCA2 genes (factors that repair damaged DNA and are strongly associated with the development of hereditary breast and ovarian cancer [HBOC]) (10). Thus, the physiological action of CagA causes an accumulation of mutations in multiple cancer-related genes within a single cell; however, the molecular mechanism underlying the development of gastric cancer caused by H. pylori infection remains unclear.

Previous studies have suggested that homologous recombination deficiency (HRD) of genes, including BRCA1 or BRCA2, may be linked to the development of gastric cancer (11-14). Therefore, our medical group compared the number of patients with gastric cancer in families with and without HBOC (n = 91 and 94, respectively) and found that families with HBOC had a greater proportion of patients with gastric cancer. Furthermore, the majority of patients with gastric cancer in HBOC families were males. Therefore, oral administration of poly-ADP-ribose-polymerase (PARP) inhibitor, which is an effective drug for patients with platinum-sensitive HBOC or ovarian cancer with HRD, may be used in patients with gastric cancer with BRCA1/2 with pathogenic variants (PVs) and/or HRD.

To confirm the efficacy of this drug, patients with advanced/metastatic gastric cancer with PVs detected in 10 genes included in the homologous recombination (HR) genes (ATM, BARD1, BRCA1, BRCA2, BRIP1, CDK12, CHEK2, PALB2, RAD51C, and RAD51D) were randomized to receive either olaparib plus paclitaxel or paclitaxel alone. We compared the safety and efficacy of the combination of olaparib and paclitaxel and paclitaxel alone as second-line chemotherapy for advanced/metastatic gastric cancer and found that the overall survival (OS) was significantly longer in the olaparib and paclitaxel combination group than in the paclitaxel group. Naturally, clinical trials with large cohorts should be conducted to further verify the efficacy of olaparib in advanced/metastatic gastric cancer with PVs, BRCA1/2, or HRD. The present study summarizes these results and suggests that new gastric cancer treatments based on our cancer gene panel testing can be considered in the future.

## Patients and methods

### 1. Cancer genome profiling

We conducted a multi-center retrospective observational analysis of patients who underwent cancer genomic medicine at cancer medical facilities in Kyoto, Japan. The study complied with the principles of the Declaration of Helsinki and was approved by the Central Ethics Review Board of the National Hospital Organization Headquarters, Tokyo, Japan (approval number: NHO R4-04; dated: November 18, 2020) and the Kyoto University School of Medicine, Kyoto, Japan (approval number: M237; dated: August 24, 2022). All participants provided written informed consent. Our clinical research complied with the Helsinki Statement.

The cancer genomic medicine is carried out using cancer gene panel testing, which was approved by the Japanese Ministry of Health, Labor and Welfare in June 2019. The following panels were tested —OncoGuide™ NCC Oncopanel gene mutation analysis set (Sysmex Corporation Kobe, Hyogo, Japan) and FoundationOne CDx’s cancer genome test (Foundation One CDx, FoundationOne liquid CDx; Foundation Medicine, Inc., Cambridge MA, USA).

### 2. Clinical trial

In this phase-II double-blind clinical trial (Study 39; NCT01063517), patients were randomly assigned to receive either oral olaparib (100 mg twice per day; tablets) plus paclitaxel (80 mg/m^2^ per day; intravenously on days 1, 8, and 15 of every 28-day cycle) or placebo plus paclitaxel (placebo/paclitaxel), followed by maintenance monotherapy with olaparib (200 mg twice per day) or placebo. The study population was enriched to 50% for patients with BRCA1/BRCA2 with PVs and/or HRD. The primary endpoint was progression-free survival (PFS) and overall survival (OS) (Supplementary Data).

### 3. Ethical considerations

#### Institutional Review Board (IRB) Approval and Consent to Participate

This research on human cancer genome information derived from results of cancer genome gene panels was conducted at the Kyoto University, its affiliated hospitals, and the National Hospital Organization Kyoto Medical Center in accordance with institutional guidelines (IRB approval no. 50-201504, NHOKMC-2023-2, and H31-cancer-2). All patients were briefed on the clinical study and agreed to take part in the present study by providing informed consent for participation. Our clinical research complied with the Helsinki Statement.

Ethics committee name: IRB of the National Hospital Organization Headquarters (approval code: H31-cancer-2; approval date: November 09, 2019, and June 17, 2013).

Ethics committee name: IRB of Kyoto University (approval code: R34005; approval date: August 01, 2022).

### 4. Ethical compliance with human study

This study involves research with human participants and was approved by the institutional ethics committee(s) and IRBs. This manuscript contains personal and/or medical information and a case report/case history about an identifiable individual; therefore, it has been sufficiently anonymized in line with our anonymization policy. The authors obtained direct consent from the patient.

The authors attended research ethics education through the Education for Research Ethics and Integrity (APRIN e-learning program (eAPRIN)) agency. The completion numbers for the authors are AP0000151756, AP0000151757, AP0000151769, and AP000351128.

### 6. Statistical analysis

All data are expressed as the mean and standard error of the mean. The normality of the data distribution was verified using the Shapiro–Wilk test; accordingly, between-group comparisons were made using the unpaired two-tailed *t*-test or the Mann–Whitney *U* test. Multiple comparisons were performed using a one-way analysis of variance with a Tukey post hoc test or a Kruskal–Wallis analysis with a post hoc Steel-Dwass or Steel test. A *p*-value of <0.05 was considered statistically significant. All statistical analyses were conducted using the JMP software (SAS Institute, Cary, NC, USA).

### 7. Data availability

The data supporting the findings of this study can be obtained from the corresponding author upon reasonable request. Details of the materials and methods are provided in the supplementary files available online. The name of the supplementary data which contains this information; Material and Methods, *Study Design, Random Assignment and Masking, Study End Points and Assessments, Statistical Analysis, Management of non-hematological treatment-related AEs attributed to Olaparib, Management of non-hematological treatment-related AEs attributed to paclitaxel*.

Details of Materials and Methods are indicated in Supplementary files, which are available online.

## Results

There is a paucity of literature on the combined effects of germline and/or somatic PVs in driver genes and H. pylori infection on the risk of gastric cancer onset and progression. It is known that H. pylori CagA injected into gastric mucosal epithelial cells significantly increases the accumulation of gene mutations that lead to gastric cancer (15) and that the CagA genotype tends to have a significant influence on the development of gastric cancer (15, 16). BRCA1/2 genes suppress tumor formation and progression, and recent studies have reported that CagA may inhibit the suppressive effect of BRCA1/2 on the development of HBOC (11-14). Therefore, this action of CagA presumably leads to the accumulation of genetic mutations in BRCA1/2 (i.e., PVs in BRCA 1/2) that trigger the onset and progression of gastric cancer by indirectly inducing cancerous transformation of gastric mucosal epithelial cells. Furthermore, there may be a high incidence of gastric cancer in patients from HBOC families with germline mutations in the BRCA1/2 genes (11-14). Therefore, we investigated the incidence of gastric cancer in patients with (n = 91) and without (n = 94) a history of HBOC (families consisting of 1 to 4 or 6 generations). We found that the HBOC families had a higher incidence of gastric cancer (3.55%) compared to the non-HBOC families (0.78%) (Table 1, S.Figure 1, S.Figure 2). Furthermore, 74.41% of the patients with gastric cancer in the HBOC families were males compared to 55% in the non-HBOC families (Table 1, S.Figure 1, S.Figure 2).

**Table 1.** Distribution of patients with hereditary gastric cancer in HBOC families reflecting the extent of genetic testing^1^.

| Generation | HBOC (n = 78 families) |  | Non-HBOC (n = 86 families) |  |
| --- | --- | --- | --- | --- |
|  | Patients with GC<sup>2</sup><br>(other tumor<sup>3</sup>) | Incidence% of GC<sup>2</sup><br>(Total people) | Patients with GC<sup>2</sup><br>(other tumor<sup>3</sup>) | Incidence (%) of GC<sup>2</sup><br>(Total people) |
| I | 15 (26) cases | 4.36% (344 cases) | 1 (6) case | 0.31% (358 cases) |
| II | 47 (197) cases | 6.18% (761 cases) | 6 (45) cases | 0.74% (730 cases) |
| III | 19 (145) cases | 3.24% (587 cases) | 11 (42) cases | 1.73% (572 cases) |
| IV | 4 (12) cases | 0.99% (405 cases) | 2 (7) cases | 0.55% (395 cases) |
| V | 1 (1) case | 0.33% (299 cases) | 0 (2) cases | 0.00% (281 cases) |
| VI | 0 (0) case | 0.00% (28 cases) | 0 (0) cases | 0.00% (41 cases) |
| Total cases | 86 (381) cases | 3.55% (2424 cases) | 20 (102) cases | 0.84% (2377 cases) |

**Sex-wise distribution of patients with gastric cancer**
| Total cases | Patients with GC in HBOC family (86 cases) |  | Patients with GC in No-HBOC family (20 cases) |  |
| --- | --- | --- | --- | --- |
|  | 64 males with GC | 74.41% males | 11 males with GC | 55.0% males with GC |

Genetic testing<sup>1</sup>: BRACAnalysis® Diagnostic System (Myriad Genetics G.K., Zurich, Switzerland); GC<sup>2</sup>: gastric cancer; other tumor<sup>3</sup>: HBOC-related cancers (hereditary cancers including breast cancer, ovarian cancer, prostate cancer, and pancreatic cancer).

From December 2019 to June 2024, a total of 36,211 new treatments were investigated using cancer genome panel tests (FoundationOne® CDx test: n = 27,981 cases; OncoGuide™ NCC oncopanel test (Riken Genesis, Yokohama, Japan): n = 8,230 cases) in cancer genomic medicine conducted at the Japanese national universities. A new treatment method was examined using cancer genome panel testing in 2361 Japanese patients with advanced/metastatic gastric cancer; of these, a total of 2340 patients (99.11%) were found to be infected with H. pylori as a result of cancer genome testing and underwent cancer genome panel testing. Recent clinical studies have reported an incidence of 5%–10% for H. pylori infection in the Japanese population. In our study, BRCA2 with germline PVs (*g*PVs) and/or somatic PVs (sPVs) were detected in 395 patients (16.73%) with advanced/metastatic gastric cancer. These findings are consistent with the histopathological findings showing reduced progression of epithelial cells in the gastric mucosal tissue of mice in a study using genetically modified Gan mice (Gan^*tgBrca2*^) (17). Further, ERBB2 with PVs was detected in 583 patients with advanced/metastatic gastric cancer. Our results in cancer genomic medicine are similar to those of a clinical study conducted by Dr. He et al. (18).

The genome panel testing revealed that BRCA2 with *g*PV and/or *s*PV was detected in 395 patients (16.73%: 395/2361) with advanced/metastatic gastric cancer. Therefore, oral administration of PARP inhibitors was started in these patients. All patients with advanced/metastatic gastric cancer (n = 132 cases) were randomly assigned to receive either olaparib plus paclitaxel (n = 67 cases) or paclitaxel alone (n = 65 cases) (Table 2); additionally, 68 patients with advanced/metastatic gastric cancer who had a PV detected in any of the 10 HRD genes were randomly assigned to receive either olaparib plus paclitaxel (n = 35 cases) or paclitaxel alone (n = 33 cases) (Table 2). To evaluate its suitability as a second-line treatment for advanced/metastatic gastric cancer, the efficacy and safety of olaparib in combination with paclitaxel and paclitaxel alone were compared. We found that the PFS was significantly longer in the olaparib plus paclitaxel group compared with the paclitaxel alone group (all cases: hazard ratio [HR] = 0.81, 95% confidence interval [CI] = 0.61–1.04, p = 0.032; gastric cancer cases with HRD: HR = 0.61, 95% CI = 0.52–1.11, p = 0.067). Likewise, OS was significantly longer in the olaparib plus paclitaxel group compared with the paclitaxel alone group (all cases: HR = 0.57, 95% CI = 0.34–0.86, p = 0.010; gastric cancer cases with HRD: HR = 0.36, 95% CI = 0.16–0.72, p = 0.003).

**Table 2.**
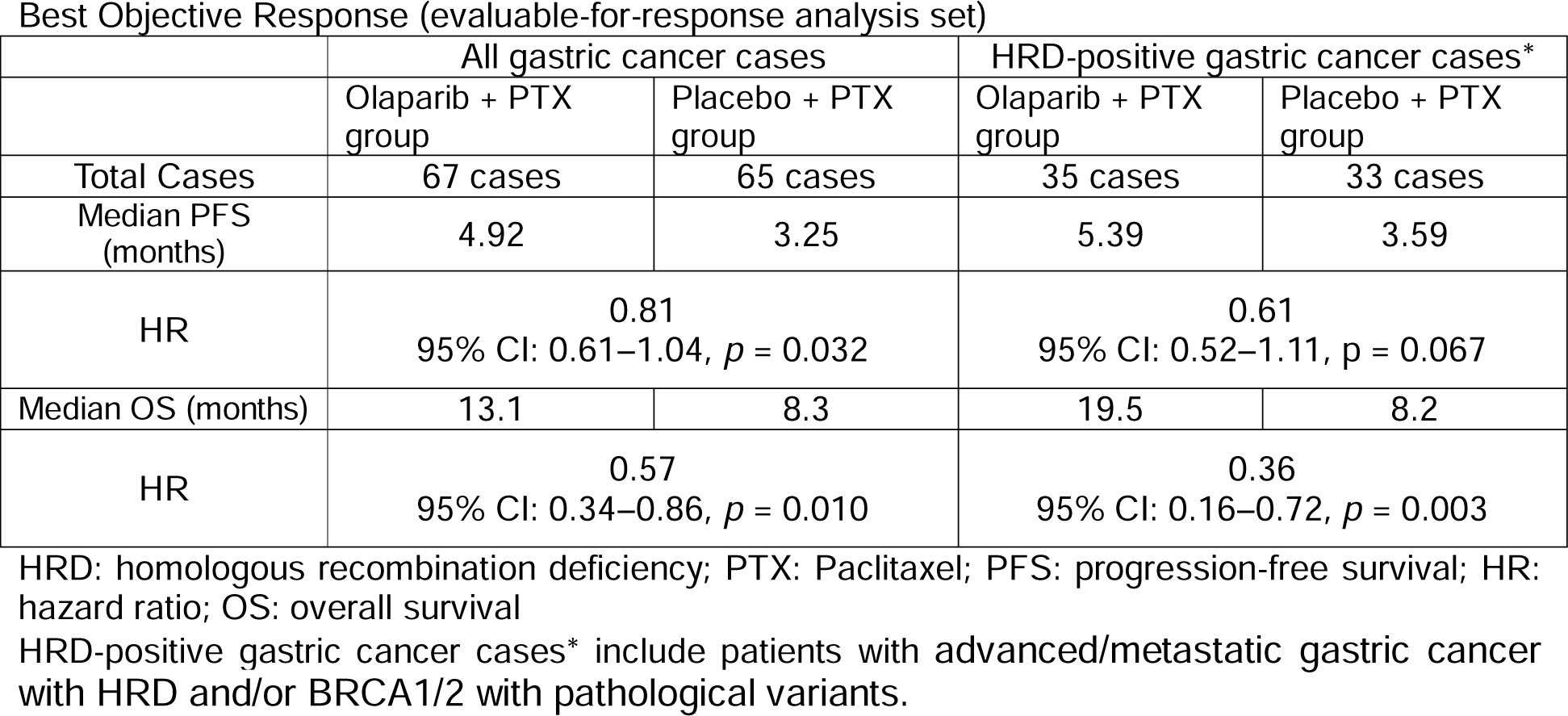
Clinical effects of the combination therapy with olaparib and paclitaxel for patients with HRD-positive gastric cancer.

Table 3 lists the most commonly observed adverse events (AEs) observed in our patients. The proportion of patients experiencing AEs was similar in both groups and corresponded to grade ≥ 3 as per the Common Terminology Criteria for AEs (all cases — olaparib/paclitaxel: n = 47, 70.1%; placebo/paclitaxel: n = 48, 73.8%; gastric cancer patients with HRDs — olaparib/paclitaxel: n = 25, 71.4%; placebo/paclitaxel: n = 23, 70.0%). A higher proportion of patients reported serious AEs (SAEs) in the placebo/paclitaxel arm than the olaparib/paclitaxel arm; pneumonia was the most common SAE (all cases — olaparib/paclitaxel: n = 4, 5.9%; placebo/paclitaxel: n = 6, 9.2%; gastric cancer patients with HRDs — olaparib/paclitaxel: n = 2, 5.7%; placebo/paclitaxel: n = 2, 6.1%). No changes of significant clinical impact were noted in any chemical parameter. The AEs observed were consistent with the known profile of paclitaxel. No unexpected safety signals or clinically important changes in vital signs, electrocardiogram parameters, or physical examination were recorded in the entire cohort during the study. No SAEs with an outcome of death that can be causally related to study treatment (according to investigator assessment) were reported.

**Table 3.** AEs (any grade) reported in 20% of patients overall or Grade ≥ 3 AEs reported in 5% of patients overall (arranged by MedDRA Preferred Term)

| AE | No. (%) |  |  |  |  |  |  |  |
| --- | --- | --- | --- | --- | --- | --- | --- | --- |
|  | All patients with gastric cancer |  |  |  | Patients with HRD-positive gastric cancer |  |  |  |
|  | Olaparib/Paclitaxel (n = 67) |  | Placebo/Paclitaxel (n = 65) |  | Olaparib/Paclitaxel (n = 35) |  | Placebo/Paclitaxel (n = 33) |  |
|  | Any Grade | Grade > 3 | Any Grade | Grade > 3 | Any Grade | Grade > 3 | Any Grade | Grade > 3 |
| Hematologic AEs |  |  |  |  |  |  |  |  |
| Anemia | 12 (18) | 7 (11) | 12 (19) | 7 (11) | 6 (17) | 3 (9) | 5 (17) | 3 (9) |
| Neutropenia | 50 (75) | 31 (46) | 40 (65) | 25 (39) | 26 (74) | 6 (17) | (70) | 7 (21) |
| Nonhematologic AEs |  |  |  |  |  |  |  |  |
| Alopecia | 31 (46) | 0 | 31 (47) | 0 | 15 (43) | 0 | 23 (40) | 0 |
| Decreased appetite | 23 (34) | 0 | 27 (42) | 0 | 11 (32) | 0 | 5 (31) | 0 |
| Neuropathy peripheral | 24 (36) | 2 (3) | 14 (21) | 1 (2) | 10 (31) | 0 | 8 (23) | 0 |
| Nausea | 21 (31) | 0 | 26 (40) | 0 | 9 (26) | 8 (22) | 7 (22) | 6 (18) |
| Asthenia | 20 (30) | 2 (3) | 18 (27) | 7 (10) | 10 (28) | 0 | 10 (20) | 0 |
| Diarrhea | 21 (31) | 2 (3) | 18 (27) | 1 (2) | 11 (29) | 2 (6) | 7 (25) | 0 |
| Abdominal pain | 15 (22) | 0 | 16 (24) | 2 (3) | 3 (21) | 0 | 1 (18) | 0 |
| Fatigue | 16 (24) | 1 (10.4) | 21 (32) | 2 (3) | 3 (23) | 3 (9) | 6 (23) | 3 (9) |
| Myalgia | 10 (15) | 0 | 23 (36) | 0 | 7 (13) | 2 (6) | 12 (36) | 3 (9) |
| Pneumonia | 4 (6) | 2 (3) | 6 (9) | 3 (5) | 2 (6) | 1 (3) | 2 (6) | 1 (3) |

AE: adverse event; MedDRA: Medical Dictionary for Regulatory Activities.
In addition to the cases of neutropenia, two patients experienced AEs of febrile neutropenia. Both events occurred in the olaparib/paclitaxel arm during the combination phase; one event was graded 3 as per the Common Terminology Criteria for Adverse Events; the other was graded 2 and led to dose modification. “Patients with HRD-positive gastric cancer” refer to patients with advanced/metastatic GC with HRD and/or BRCA1/2 with PVs.

## Discussion

The American Cancer Society reports that an estimated 24,590 Americans will be diagnosed with gastric cancer each year, and an estimated 10,720 Americans will die from the disease (19). The five-year survival rate for gastric cancer (all stages combined) is about 28% (19). BRCA1 and BRCA2 are proteins that repair DNA damage and are expressed in all cells, i.e., these proteins work to suppress the neoplastic transformation of normal cells. Accordingly, a link between HBOC and the development of gastric cancer has been pointed out; however, the detailed mechanism by which H. pylori infection induces the development of gastric cancer remains unclear. Using the information on hereditary cancers gained based on cancer genomic medicine conducted at university hospitals in Japan, our research has revealed a high incidence of gastric cancer in HBOC families, with the incidence in HBOC families being slightly higher in men than in women. Furthermore, we found that treatment with PARP inhibitors may be effective in such patients with hereditary gastric cancer.

In the United States, the incidence of gastric cancer is relatively low compared to other types of cancer; however, globally, gastric cancer is the second most common cause of cancer-related mortality. Gastric cancer is quite prevalent in Asian countries, such as Korea, China, Taiwan, and Japan, and its treatment usually involves surgical removal of the cancer, followed by chemotherapy, with or without radiation therapy. In Japan, gastric cancer is associated with more than 50,000 deaths annually (20). H. pylori is a common microbe found in East Asia, including Japan (20); as a result, it is believed that many patients in East Asia develop gastric cancer because of H. pylori infection. The bacterium H. pylori is broadly classified into CagA-positive and CagA-negative strains based on the presence or absence of CagA secretion, with the CagA-positive strains inducing relatively more severe gastric mucosal lesions (20-23). In Western countries, the ratio of CagA-positive to CagA-negative strains is approximately 6:4 (21); however, CagA-positive H. pylori strains are more common in East Asia, including Japan, are. Consequently, the incidence of gastric cancer in the Japanese population (gastric cancer characteristic of East Asia) is significantly higher (males = 6.07%; females = 2.11%) than in Western countries (USA: males = 0.65%, females = 0.33%; UK: males = 0.66%, females = 0.28%) (24).

H. pylori infects both men and women, with no significant differences in the infection rate between the two sexes. However, compared to non-infected people, the probability of developing gastric cancer from birth to 85 years of age in people with H. pylori infection is 17.0% (about 1 in 6 people) for males and 7.7% (about 1 in 13 people) for females. This indicates that the incidence of gastric cancer due to H. pylori infection is higher in males than in females. Presumably, these sex-related differences observed in our study were due to the relatively larger number of patients with gastric cancer (74.41% men) in the 91 HBOC families compared to 55% males with gastric cancer in the 94 non-HBOC families (Table 1). Nevertheless, the reason for sex-based differences in the incidence of gastric cancer in HBOC families is unclear. Further clinical studies must be conducted to clarify these reasons.

A Korean research group demonstrated that cell lines for gastric cancer, especially those with significantly low levels of ataxia telangiectasia mutations (ATM), a key activating gene in the HR gene family, were sensitive to PARP inhibitors, such as olaparib, niraparib, and talazoparib) (25). The research group compared the efficacy of the combination of olaparib and paclitaxel (olaparib/paclitaxel) with paclitaxel alone (placebo/paclitaxel) in patients with advanced/metastatic gastric cancer and evaluated whether low ATM expression was a predictor of improved clinical outcomes with olaparib/paclitaxel. They found that the combination therapy was more effective in the treatment of advanced/metastatic gastric cancer and is expected to extend the OS in patients with low ATM (25).

Another Japanese research group demonstrated that germline pathogenic mutations in nine genes of the HR gene family (APC, ATM, BRCA1, BRCA2, CDH1, MLH1, MSH2, MSH6, and PALB2) were associated with the risk of gastric cancer in Japanese patients treated at the Aichi Cancer Center Hospital Epidemiology Research Program (HERPACC), Nagoya, Achi, Japan (26, 27). They found a significant interaction between H. pylori infection and these pathogenic mutations in the HR gene regarding the risk of advanced/metastatic gastric cancer.

The HBOC syndrome is a cancer susceptibility syndrome caused by germline mutations in the BRCA1 or BRCA2 proteins and is inherited in an autosomal dominant manner (28). In addition to the HBOC causative genes, studies have identified other candidate genes that specifically contribute to the onset of HBO, i.e., genes involved in the DNA damage checkpoint ATM and CHEK2, and genes involved in the repair of DNA double-strand breaks, such as PALB2, BRIP1, and RAD51C, are reported to contribute to the onset of HBOC (29-33). Previous clinical studies have shown that PVs in these genes are closely associated with the development of breast cancer, ovarian cancer, prostate cancer, or pancreatic cancer. Furthermore, PALB2 mutation carriers carry a 35% cumulative risk of breast cancer at the age of 70 years (34), and ATM with PVs have been detected in 1.2% to 6.9% of all breast cancer cases (35).

Based on the results of previous clinical studies, ten genes in the HR gene family, which are associated with the development of breast cancer, ovarian cancer, prostate cancer, and pancreatic cancer, were considered to be deeply involved in the development of gastric cancer. Therefore, our research team conducted a comparative study of the incidence of gastric cancer in patients belonging to HBOC versus non-HBOC families and found the incidence to be greater in the HBOC families (3.55% versus 0.78%) (Table 1, S.Figure 1, S.Figure 2). PARP inhibitors are known to be effective against ovarian, breast, prostate, and pancreatic cancers in which HRD or BRCA1/2 with PVs have been detected. Therefore, all patients with gastric cancer were randomly assigned to receive either olaparib plus paclitaxel (n = 67 cases) or paclitaxel alone (n = 65 cases). Further, patients with gastric cancer in HBOC families were randomly assigned to receive olaparib plus paclitaxel (n = 35 cases) or paclitaxel alone (n = 33 cases). Prolonged PFS and OS were observed in gastric cancer patients from HBOC families who received the combination therapy compared to the whole study cohort. However, a previous clinical study failed to demonstrate significant improvements in the OS with olaparib in the overall or ATM-negative population of Asian patients with advanced gastric cancer (36, 37). Therefore, the combination therapy of olaparib with paclitaxel may be used as a second-line treatment for advanced/recurrent gastric cancer with HRD and/or BRCA1/2 with PVs. Further clinical trials are required to establish the regimen for combination therapy with olaparib and paclitaxel in this population.

The Japanese Ministry of Health, Labor, and Welfare has approved insurance coverage for the oral administration of PARP inhibitors — olaparib for breast, ovarian, pancreatic, and prostate cancer in patients with HRD or BRCA1/2 with PVs and niraparib for ovarian cancer with HRD or BRCA1/2 with PVs); the suitability is determined by the MyChoice test (Myriad Genetics, Inc. CA). Additionally, the has approved insurance coverage for oral administration of talazoparib for breast cancer with BRCA1/2 with PVs detected by BRACanalysis and prostate cancer with BRCA1/2 with PVs detected by FoundationOne CDx. Based on the available clinical evidence, oral administration of PARP inhibitors may be established as a treatment for gastric cancer with HRD and/or BRCA1/2 with PVs. Our research findings offer substantial evidence for guiding the establishment of early treatment for patients with advanced/metastatic gastric cancer in whom BRCA1/2 with PVs has been detected. This study generated informative efficacy and safety data regarding the use of olaparib in combination with a chemotherapeutic agent; paclitaxel and provides a foundation for future studies in this challenging patient population. The clinical trials conducted to date have included cohorts with a small number of subjects; further studies with larger cohorts are required to confirm the effectiveness of PARP inhibitors in advanced/metastatic gastric cancer with HRD and/or BRCA1/2 with PVs.

## Supporting information

HBOC

Non-HBOC

## Contributors

TH, KS, MO, and MM were involved in study design, data collection, reviewing and interpreting the data, and writing the manuscript. TH, KS, and OM was involved in the literature search, study design, data collection, data interpretation, and manuscript writing. TH, KS, and OM was involved in data collection and interpretation. TH, and OM was involved in data collection and interpretation and writing the manuscript. TH, KS, and OM, MM was involved in study conception and design, data analysis and interpretation, and writing the manuscript. TH, and OM was involved in patient recruitment, data analysis and interpretation, and writing the manuscript. TH, and IK was the medical lead for AstraZeneca for the study and participated in data collection and evaluation, as well as writing and editing the manuscript. TH, and IK was the lead physician for the study and was involved in study design and conduct, data analysis and interpretation, and manuscript review.

## Funding

This clinical research was performed with research funding from the following: Japan Society for Promoting Science for TH (grant No. 19K09840), START-program Japan Science and Technology Agency (JST) for TH (grant No. STSC20001), and the National Hospital Organization Multicenter clinical study for TH (grant No. 2019-Cancer in general-02), and The Japan Agency for Medical Research and Development (AMED) (grant No. 22ym0126802j0001), Tokyo, Japan.

### Institutional Review Board Statement

This study was reviewed and approved by the Central Ethics Review Board of the National Hospital Organization of Japan (Meguro, Tokyo, Japan) and the Central Ethics Review Board of Kyoto University Hospital (Kyoto, Kyoto, Japan). The approved number for this study is NHOKMC-2023-2 and 50-201504. In order to carry out this research, the authors attended a research ethics education course (e-APRIN) conducted by Association for the Promotion of Research Integrity (APRIN; Shinjuku, Tokyo, Japan). The approved numbers of e-APRIN are AP0000151756, AP0000151757, AP0000151758, AP0000151769.

### Informed Consent Statement

Informed consent was obtained from all subjects involved in the study. Written informed consent has been obtained from the patient(s) to publish this paper.

## Data Availability Statement

Data are available on various websites and have also been made publicly available (more information can be found in the first paragraph of the Results section). The transparency document associated with this article can be found in the online version at https://kyoto.hosp.go.jp/html/guide/medicalinfo/clinicalresearch/expand/gan.html (accessed on 18 May 2024).

## Acknowledgments

The authors appreciate Zhang W (Roche Tissue Diagnostics, Tucson, AZ, USA) for critical research assistance. The authors would like to thank Yoshimi Fujikura at Medical Information Center of AstraZeneca Inc. for clinical research information. The authors thank all medical staff for clinical research at Kyoto University School of Medicine and the National Hospital Organization Kyoto Medical Center. Conflicts of Interest The authors declare no potential conflicts of interest. The funders had no role in study design, data collection and analysis, decision to publish, or preparation of the manuscript.

## Abbreviations

AE: Adverse Event
BRCA: breast cancer susceptibility gene
CagA: Cytotoxin-associated gene A antigen
GC: gastric cancer
HBOC: hereditary breast and ovarian cancer
H. pylori: Helicobacter pylori
HRD: deficiency of homologous recombination
OS: over survival
PFS: Progression Free Survival
PARP: poly ADP-ribose polymerase
WHO: World Health Organization

