## Supplementary for "Efficacy and Tolerability of Olaparib Plus Paclitaxel in Patients with hereditary gastric cancer linked to a family history of hereditary breast and ovarian cancer (HBOC)"

### Material and Methods

#### *Study Design*

In the initial combination phase of this prospective, randomized, double-blind, multicenter clinical study (approved number; 50-201504), patients received olaparib (100 mg twice per day, continuous; tablet formulation) or matching placebo, in combination with paclitaxel (80 mg/m2 per day intravenously on days 1, 8, and 15) in 4-week treatment cycles. Patients were expected to receive six to 10 paclitaxel treatment cycles. After completing paclitaxel treatment, patients entered the maintenance therapy phase, where they received olaparib (200mgtwice per day) or placebo monotherapy until objective progression or provided that they were benefiting from treatment and did not meet discontinuation criteria. Management of toxicities by olaparib and/or paclitaxel dose modifications (reductions and/or interruptions [delays]) is described in the Data Supplement. The trial protocol was reviewed and approved by the institutional review boards of the participating institutions. The study was performed in accordance with the Declaration of Helsinki, Good Clinical Practice, and the AstraZeneca Policy on Bioethics. 28

#### *Random Assignment and Masking*

Before starting the combination phase, patients were randomly assigned 1:1 to receive olaparib/paclitaxel or placebo plus paclitaxel (placebo/paclitaxel). The random assignment scheme was produced by computer software program (GRand; AstraZeneca Global Randomization system) that generated random numbers. Blocked random assignment was generated, with all centers using the same list to minimize imbalances in numbers of patients assigned to each arm. Random assignment was distinguished by Breast Cancer. Susceptibility Gene1 (BRCA1) or/and BRCA2 with Pathogenic Variants (PVs) and/or Homological Recombination Deficiency (HRD) according to the results obtained from cancer genome testing ensuring that the proportion of patients with BRCA1/2 PVs and/or HRD in each arm was approximately 50%.

#### *Study End Points and Assessments*

The primary end point was investigator-assessed progression-free survival (PFS), defined as time to objective disease progression determined by Response Evaluation Criteria in Solid Tumors (RECIST) version 1.1 or death; PFS was analyzed in the population of all patients with gastric cancers and the population of patients with gastric cancer with BRCA1/2 PVs and/or HRD. Secondary end points were over survival (OS), objective response rate (ORR), percent change in tumor size at week 8 (all assessed in the population of all patients with gastric cancers and population of patients with gastric cancer with BRCA1/2 PVs and/or HRD), and safety/tolerability. Tumor assessment was performed at screening, every 8 weeks until week 40, and every 16 weeks thereafter, until objective disease progression as determined by the investigator. RECIST assessments were performed using computed tomography and magnetic resonance imaging scans of the chest, abdomen, and pelvis. Patients who had discontinued all study treatment and showed disease progression were observed for survival at 8-week intervals. Duration of follow-up was defined as the number of days from the time of random assignment to the date of death or data cutoff (June 09, 2024) in the absence of death for censored patients. Adverse events (AEs) and laboratory parameters were recorded throughout the trial and graded according to the National Cancer Institute Common Terminology Criteria for Adverse Events version 3.0.

#### *Statistical Analysis*

This trial was sized using one-sided 10% significance levels; an outcome of *P*_.1 (one-sided) for either the population of all patients with gastric cancers and the population of patients with gastric cancer with BRCA1/2 PVs and/or HRD would be regarded as promising (but not definitive). The study aimed to randomly assign 200 patients, of whom 68 were to be the patients with gastric cancer with BRCA1/2 with PVs and/or HRD. At the time of study design, the screening prevalence for tumor samples obtained from the patients with gastric cancer with BRCA1/2 with PVs and/or HRD was anticipated to be 19% to 24%; therefore, once 132 patients with gastric cancer without BRCA1/2 with PVs and/or HRD had been enrolled, recruitment would be restricted to the patients with gastric cancer without BRCA1/2 PVs and/or HRD only. PFS analysis was to be performed after approximately 99 progression events (approximately 50 events in the population of patients with gastric cancer with BRCA1/2 with PVs and/or HRD) and was calculated to have approximately 80% power to detect a true hazard ratio (HR) of 0.81 in the population of patients with gastric cancer without BRCA1/2 with PVs and/or HRD (0.61 in the population of patients with gastric cancer with BRCA1/2 with PVs and/or HRD), based on a one-sided 10% significance level. Efficacy was analyzed on an intent-to-treat basis in all randomly assigned patients (full analysis set), except ORR and change in tumor size, which were analyzed using an evaluable-for-response population that excluded patients.

#### *Management of non-hematological treatment-related AEs attributed to olaparib*

Non-hematological grade $\geq$3 AEs attributable to olaparib were to be managed by dose interruption, with the duration of interruption not to exceed 28 days. Before treatment could be resumed with a 50% reduction of dose, the toxicity had to recover to either 4 grade $\leq$1 or the baseline grade. If resolution of the AE did not occur within 28 days, the patient was to be withdrawn from the study.

#### Management of non-hematological treatment-related AEs attributed to paclitaxel

In the event of elevated AST (5–10xULN) and/or bilirubin (1·6–2·5 mg/dL) levels, administration of paclitaxel was to be interrupted (maximum interruption, 28 days) until the toxicity had resolved to either grade $\leq$1 or the baseline grade. Treatment with paclitaxel could then be resumed at a dose of 65 mg/m$^2$; however, the dose level could be returned to the starting dose (80 mg/m$^2$) in subsequent treatment cycles. In the event of grade 3 neuropathy, paclitaxel was to be withheld until resolution to grade ≤1, and then resumed at a dose of 65 mg/m$^2$. If grade ≥3 neuropathy lasted for ≥4 weeks or occurred following dose reduction, paclitaxel treatment was to be permanently discontinued. Paclitaxel was also to be withdrawn in the event of severe hypersensitivity reaction, grade ≥3 AST/ALT levels lasting ≥7 days, or grade ≥3 bilirubin levels. For grade 3 non-hematological toxicities other than those mentioned above (and excluding nausea, vomiting, and asthenia), patients were to have a permanent dose reduction to 65 mg/m$^2$. Patients experiencing grade 4 non-hematological toxicity could have a paclitaxel dose interruption for ≥4 weeks (one treatment cycle) to permit recovery to grade ≤3, followed by a permanent dose reduction to 65 mg/m$^2$ .
